# Insomnia symptoms and risk of bloodstream infections: prospective data from the prospective population-based HUNT Study, Norway

**DOI:** 10.1101/2022.05.05.22274704

**Authors:** Marianne S. Thorkildsen, Lars E. Laugsand, Tom I.L. Nilsen, Randi M. Mohus, Lise H. Høvik, Tormod Rogne, Erik Solligård, Jan K. Damås, Lise T. Gustad

## Abstract

**Objective:** Previous research suggest decreased immune function and increased risk of infections in persons with insomnia. We examined the effect of insomnia symptoms on risk of bloodstream infections (BSI) and BSI-related mortality in a population-based prospective study.

**Methods:** A total of 53,536 participants in the Norwegian HUNT2 study (1995–97) were linked to prospective data on clinically relevant BSIs until 2011. In Cox regression, we estimated hazard ratios (HRs) with 95% confidence interval (CI) for a first-time BSI and for BSI related mortality (BSI registered ≤30 days prior to death) associated with insomnia symptoms.

**Results:** Compared with participants who reported “no symptoms of insomnia”, participants reporting having “difficulty initiating sleep” often/almost every night had a HR for a first-time BSI of 1.14 (95% CI 0.96–1.34). Participants reporting “difficulties maintaining sleep” often/almost every night had a HR of 1.19 (95% CI 1.01–1.40), whereas those having a “feeling of non-restorative sleep” once a week or more had a HR of 1.23 (95% CI 1.04–1.46). Participants experiencing all three above insomnia symptoms frequently had a HR of 1.39 (1.04–1.87) and being troubled by insomnia to a degree that affected work performance was associated with a HR of 1.41 (95% CI 1.08–1.84). The HRs for BSI related mortality suggest an increased risk with increasing insomnia symptoms, but confidence intervals are wide and inconclusive.

**Conclusions:** We found that frequent insomnia symptoms and insomnia symptoms that affected work performance was associated with a weak positive increased risk of BSI.

## Introduction

Insomnia is a sleep disorder defined as having a subjective feeling of difficulties initiating or maintaining sleep, or early morning awakening and impaired daytime functioning. Insomnia affects from 6–19% of the European population (Riemann et al., 2017), and women have a higher predisposition for insomnia than men (Zhang & Wing, 2006). Sleep is a restorative process important for the immune system and insomnia can be detrimental to both the innate and adaptive immune system (Besedovsky, Lange, & Haack, 2019a) and affect genes for immune functioning (M. R. Irwin, 2019). Biological mechanisms related to the effect of insomnia on immune responses include elevated C-reactive protein levels (Meier-Ewert et al., 2004), decreased natural killer cell activity (M. Irwin et al., 1994), suppressed interleukin 2 (IL2) and interleukin 6 (IL6) response (Burgos et al., 2006; Vgontzas, Liao, Bixler, Chrousos, & Vela-Bueno, 2009), decreased T-cell cytokine production (M. Irwin et al., 1996), reduced vaccination response (Lange, Perras, Fehm, & Born, 2003) and impaired performance of the pro-inflammatory cytokine genes IL-6 mRNA and TNFα mRNA (M. R. Irwin, Wang, Campomayor, Collado-Hidalgo, & Cole, 2006).

Previous studies investigating the association of sleep with risk of infections have mainly focused on risk of respiratory infections (Cohen, Doyle, Alper, Janicki-Deverts, & Turner, 2009; Lin, Liu, Chung, & Chien, 2018; Patel et al., 2012; Prather, Janicki-Deverts, Hall, & Cohen, 2015; Prather & Leung, 2016) and report that participants with sleep disorders and short sleep duration were more susceptible to respiratory infections especially in the younger adult age groups (Mackenzie, 2016). Whether insomnia is associated with the risk of other and more severe invasive infections, such as bloodstream infections (BSIs) is not clear. BSI is a serious infectious disease, rating among top seven causes of deaths in Europe and North America (Goto & Al-Hasan, 2013), men are more at risk (Randi Marie Mohus et al., 2021), and BSI often leads to sepsis (Laupland et al., 2005). Sepsis is defined as a dysregulated host response to infection, estimated to annually afflict 48.9 million people and cause 11 million deaths worldwide (Rudd et al., 2020). Insomnia is thought to affect risk of all-cause mortality and cardiovascular disease mortality (Pienaar et al., 2021), however we are not aware of studies assessing if insomnia symptoms are associated with BSI risk and BSI related mortality.

In this study, we prospectively examined if insomnia symptoms was associated with the risk of clinically relevant BSI with verified microbes (Mehl et al., 2017) as well as with risk of BSI related mortality using data from the population-based HUNT study in Norway. The microbiology data allowed us to also examine the association of insomnia and risk of specific bacterial BSIs, such as *Staphylococcus (S) aureus, Escherichia (E) coli* and *Streptococcus (S) pneumonia*.

## Method

### Study population

This study is based on data from the second HUNT study in Norway (HUNT2) conducted in 1995–97. All inhabitants ≥20 years (n=93,989) in the geographical region of Nord-Trøndelag were invited to participate, and 65,237 (69.5%) attended the survey, filled in extensive questionnaires and met for a clinical examination (Krokstad et al., 2013). Of these, 1,188 participants were excluded due to either having a first BSI event, death or migration before start of follow-up (for details see outcome ascertainment). Moreover, 10,513 (16.1%) participants did not answer any of the insomnia questions, leaving a total of 53,536 persons in the analytical sample (Figure 1).

### Outcome ascertainment

HUNT2 data was linked to the hospital-based sepsis registry in Nord-Trøndelag Hospital Trust (HNT HF) by the unique 11-digit Norwegian identification number. The HNT HF sepsis registry prospectively collects information on all bloodstream infections (BSIs) that require hospitalization at the two community hospitals, Levanger Hospital (from 1994) and Namsos Hospital (from September 1999) or at the tertiary referral hospital St. Olavs hospital, Trondheim (from 1994). We defined our outcome variables as first-time BSI, and first-time BSI by the most common bacteria; *E. coli, S. aureus and S. pneumoniae*. Our definition of BSI related mortality was a BSI registered ≤30 days prior to death, as this is previously established as the gold standard outcome for deaths from BSIs requiring hospital stays (Laupland et al., 2019). For persons with multiple positive blood cultures, a new episode of BSI was defined as a positive blood culture more than 30 days after the previous episode. Isolates only consisting of Coagulase negative *Staphylococcus species, Corynebacterium species* and *Cutibacterium* species were not considered as BSI, as these bacteria are associated with contamination of blood cultures with bacterial skin flora (Paulsen et al., 2017). The participants were followed up until they experienced an event or until censoring by death, emigration, or end of follow up December 31^st^, 2011.

### Insomnia

The HUNT insomnia questionnaire included three items. One question was related to difficulty initiating sleep (“Have you had difficulties falling asleep in the last month”) and another related to difficulty maintaining sleep (“During the last month, have you woken up too early and not been able to get back to sleep”). Both questions had the response options never (0 points), occasionally (1 point), often (2 points), and almost every night (3 points). The third question was related to having a feeling of non-restorative sleep (“How often do you suffer from poor sleep”), with the response option never or few times a year (0 points), one to two times per month (1 point), about once a week (two points), more than once a week (three points). For each item, zero points were defined as the reference (no insomnia symptoms), one point as the middle value, and two or three points were merged into the highest category (frequent insomnia symptoms). The first two questions were asked to all participants 20 years and older (n=53,536), whereas the last question was only asked participants between 20–69 years (n= 44,271). We summarized into a cumulative insomnia symptom score based on the score on the three above insomnia items. Each item gave zero points to the cumulative insomnia score if no symptoms where present, else it gave 1 point. Cumulative insomnia thus ranged from 0 to 3 insomnia items present. Calculation of the cumulative insomnia score was only performed among those without any missing on each insomnia item, as only participants below 70 were asked if they had a feeling of non-restorative sleep.

Participants aged 20–69 years (n=44,271) were also asked if symptoms related to sleep influenced their work performance (“During the last year, have you been troubled by insomnia to such a degree that it influenced your work performance”, with the response option “yes” or “no”).

### Covariates

#### Demographic factors

Age and sex were obtained from the National Population Registry at the time of HUNT2 participation. Other socio-demographic factors were self-reported. We categorized marital status into never married, married, separated/widower/divorced (Hauan, Strand, & Laugsand, 2018). Education level was categorized as <10 years, 10–12 years and >12 years of fulfilled schooling (Askim et al., 2018). Participants answering “yes” to being employed, self-employed or working fulltime in the home was categorized as working. If they answered “yes” to being retired or unemployed, they were categorized as not working. Participants answered “yes” or “no” to whether they worked shift, night or on call.

#### Clinical information

Weight (kg) and height (cm) was measured by trained nurses while the participants wore light clothes and no shoes and rounded to the closest half kilo and centimeters (Holmen et al., 2003). Body mass index (BMI) was then calculated from weight in kilos (kg) divided by the squared value of height (m^2^) and categorized according to World Health Organization (WHO) recommendations; <18.5, 18.5–24.9, 25–29.9, 30.0–34.9, 35.0–39.9 and ≥40.0 kg/m^2^ (R. M. Mohus et al., 2018).

#### Lifestyle factors

Weekly alcohol consumption was based on participants’ report of how many units of beer, wine and liquor they usually consumed in a two-week period. The amount of beer, wine and liquor units was combined and categorized as “abstainer” (zero units per week or answering yes to being an abstainer), “light drinker” (1–14 units), “moderate drinker” (15–28 units) and “heavy drinker” (>28 units) (Paulsen et al., 2017). A total of 10% of the participants did not answer to the questions related to alcohol intake. Some of these participants did answer how many days a month they on average consumed alcohol, and this information was added to reduce missing data. Answering “zero times a month” was included in the “abstainer” group, “one-three times a month” in “light drinker”, “four-seven times a month” as “moderate drinker” and “more than seven times a month” as “heavy drinker”.

Smoking habits was categorized as never, former, or current smoker. In addition, physical activity was categorized into four groups based on how many hours of light and hard activity they performed. Light activity was defined as activity not involving sweating or feeling of breathlessness. Participants performing neither hard nor light activity were categorized as inactive, less than three hours of light activity and no hard activity as light activity, three or more hours of light or less than three hours of hard activity as moderate activity, and more than three hours of hard activity as vigorous activity (Paulsen et al., 2017).

#### Chronic somatic disorders

Self-reported information regarding comorbid chronic disorders was summarized into one variable answering yes/no to having at least one condition. Cardiovascular disease (CVD) was defined as having a history of acute myocardial infarction, angina pectoris, stroke, or surgery for intermittent claudication. Participants answering “yes” to having had productive cough continuously for more than three months every year for the last two years were defined as having lung disease. Cancer and diabetes diagnosis were defined by participants answering “yes” to having/or having had a diagnosis. Rheumatoid disease was defined as answering “yes” to having ankylosis spondylitis and/or rheumatoid arthritis. Participants with missing information on each of the comorbid conditions were categorized as not having the specific comorbid conditions.

#### Sleeping medication and circadian factors

As the HUNT survey is close to the Arctic Circle, being examined in the months April through August was categorized as high light exposure, from October through February as low light exposure and March and September as neither high nor low light exposure (Sivertsen, Krokstad, Overland, & Mykletun, 2009). We defined chronic use of sedatives as using sedative one or more times a week.

#### Depression and anxiety symptoms

Depression and anxiety symptom levels were self-reported and assessed using the hospital anxiety and depression scale (HADS) (Zigmond & Snaith, 1983). The scale consists of 14 items, seven questions related to depression symptoms and seven related to anxiety symptoms. The scale on each scale range is from 0–21, with increasing scores indicating increasing symptom load. Per protocol, we replaced 1–2 missing items on the HADS depression and HADS anxiety subscale with 6/7 and 5/7 of the values provided (Askim et al., 2018) and defined these as our complete case.

#### Laboratory measurements

Non-fasting serum blood samples was drawn from each HUNT participant by trained study nurses and analyzed at the Central Laboratory at Levanger Hospital. Creatinine levels were measured using Jaffe method with sample blank (Roche Diagnostics, Mannheim, Germany). Chronic kidney disease (CKD) was defined as having an eGFR <60 ml/min per 1.73 m^2^. The eGFR was estimated using the Modification of Diet in Renal Disease (MDRD)-formula, from recalibrated creatinine values (Hallan, Astor, & Lydersen, 2006).

### Statistical analysis

We imputed missing data using multiple imputation by fully conditional specification generating a total of 10 complete datasets in the “mi chained”-routine and compared these with results from complete case. Information on 12 variables from demography, social status, health behaviors, mental health and time to event were used as predictor variables to ensure the required assumption of ‘missing at random. Cox regression was used to estimate the hazard ratio (HR) with 95% confidence interval (CI) for first-time BSI, first-time BSI stratified by specific bacteria, and BSI related mortality associated with each of the insomnia symptoms and cumulative insomnia symptoms. For all comparisons, participants with no insomnia complaints were used as the reference group. All estimated effects were adjusted for first age (as the time scale), which constitutes the crude analysis. Thereafter we added adjustments for sex, marital status, education, physical activity, alcohol consumption, BMI, and smoking habits. Factors that could be mediators of the association between insomnia and risk of BSI, such as common chronic disorders, depression or anxiety symptoms were adjusted for in sensitivity analyses. Among the 44,271 participants with employment, we evaluated whether sleep problems influencing work performance was associated with risk of BSI after additional adjustment for shiftwork. Possible linear trend across insomnia categories were assessed in analysis where the insomnia categories (0–3) were entered as continuous variables in the regression model. The proportional hazards assumptions were evaluated by Schoenfeld residuals and visual inspection of log-log plots. Censoring information on death or migration was obtained from the National Population Register. As risk of infection and BSI varies by age and sex, possible effect modification by age (±60 years) and sex (man, woman) was evaluated in a likelihood ratio test of a product term of insomnia and age, and of insomnia and sex.

We performed several sensitivity analyses to address the robustness of our findings. To reduce possible influence by reverse causation we excluded the first five years of follow-up. We evaluated whether varying light exposure from seasonal changes influenced our results by stratifying the population by circadian light exposure on time of insomnia assessment in HUNT2. We stratified those who reported insomnia symptoms by months with daylight predominance. Lastly, the potential effect of use of sedatives was examined by assessing the risk of first-time BSI only among those who reported no use of sedatives. The statistical analyses were performed using STATA version 16 (Statcorp, Texas, USA).

### Ethics

Participation in the HUNT2 survey is voluntary, and participants are always able to withdraw from the health survey by contacting the HUNT research center. All information will then be deleted, and blood and urine samples annihilated. All participants signed a written consent to data collection and to linkage of data to other registers. This study has been approved by the Regional ethical committee for medical research (REK 2018/1819/REKmidt and REK 2012/153), and by the HUNT data access committee. From the data access committee in HNT HF we have approval to use and merge the HUNT data file with the HNT sepsis registry.

## Results

During a median follow-up of 14.9 years, 1579 (3.0%) of the participants experienced an episode of BSI and 284 (0.5%) suffered from BSI related mortality. Baseline characteristics of the participants are shown in Table 1.

**Table 1.** Baseline characteristics of the study population at inclusion in HUNT2

|  |  |  |  |  |
| --- | --- | --- | --- | --- |
|  |  | **By difficulties initiating sleep** |  |  |
|  | **Missing** | **Never** | **Occasionally** | **Often/almost every night** |
| **Sex** (female) | 0 | 14,601 (50.3) | 11,128 (58.0) | 3,101 (66.7) |
| **Smoking** (current) | 975 | 7,046 (24.7) | 5,667 (30.1) | 1,762 (38.8) |
| **Marital status** (married) | 114 | 18,163 (62.6) | 11,651 (60.8) | 2,690 (58.0) |
| **Education** (10-12 years) | 2,370 | 12,616 (45.2) | 7,785 (42.5) | 1,592 (36.8) |
| **Alcohol consumption** (light) | 1,618 | 13,906 (49.2) | 8,639 (46.5) | 1,729 (38.8) |
| **Physical activity** (light) | 5,159 | 7,729 (29.2) | 5,619 (32.5) | 1,468 (37.0) |
| **Working shifts** (yes) | 16,342 | 4,255 (20.2) | 2,842 (22.1) | 577 (22.2) |
| **Comorbidity** |  | 6,526 (80.9) | 1,189 (13.2) | 460 (5.1) |
| CVD | 32 | 1,812 (6.2) | 1,602 (8.4) | 653 (14.1) |
| CKD | 113 | 1,008 (3.5) | 863 (4.5) | 310 (6.7) |
| Lung disease | 282 | 2,653 (9.2) | 2,096 (11.0) | 736 (16.0) |
| Cancer | 2,651 | 897 (3.2) | 769 (4.3) | 272 (6.5) |
| Diabetes | 98 | 757 (2.6) | 576 (3.0) | 212 (4.6) |
| Rheumatic disease | 4,875 | 916 (3.4) | 874 (5.1) | 325 (8.5) |
| **Use of sleep medicine weekly** | 5,270 | 590 (2.3) | 1,448 (8.3) | 1,721 (40.6) |
| **Age (SD)** | 0 | 48.4 (16.7) | 51.5 (16.9) | 55.5 (17.3) |
| **BMI kg/m<sup>2</sup> (SD)** | 291 | 26.3 (4.0) | 26.4 (4.0) | 26.6 (4.4) |
| **Depression Score (SD)** | 1,484 | 2.8 (2.7) | 3.9 (3.0) | 5.6 (3.8) |
| **Anxiety score (SD)** | 2,260 | 3.4 (2.8) | 4.9 (3.3) | 7.0 (4.3) |
| Abbreviations: BMI=body mass index, CVD=cardiovascular disease, CKD=chronic kidney disease. |  |  |  |  |

HR for first-time BSI associated with each of the insomnia items and cumulative insomnia symptom score are presented in Table 2. Compared with participants reporting “no symptoms of insomnia”, participants experiencing “Difficulties initiating sleep” often/almost every night had a HR of 1.14 (95% CI 0.96–1.34); and “Difficulties maintaining sleep” had a HR of 1.19 (95% CI 1.01–1.40) for BSI. Participants “Having a feeling of non-restorative sleep” once a week or more had a HR of 1.23 (95% CI 1.04–1.46). Having all three cumulative insomnia symptoms was associated with a HR of 1.39 (95% CI 1.04–1.87) for BSI compared to having none of the insomnia symptoms. Moreover, the employed that responded yes to “being troubled by insomnia to such a degree that it affected work performance” had a HR of 1.41 (95% CI 1.08–1.84) for BSI, compared to the employed whom responded no to the question (Table 3).

**Table 2.**
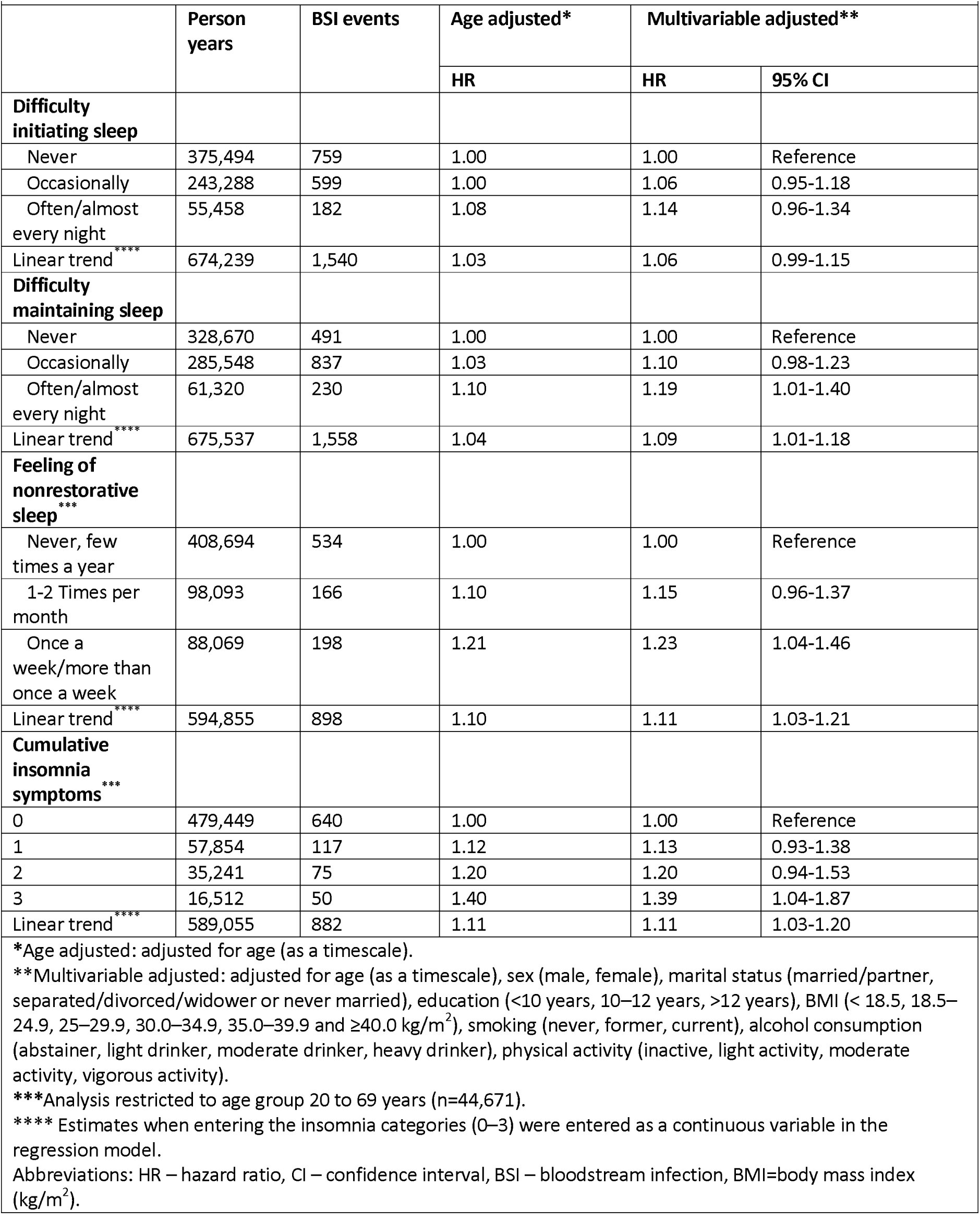
Risk of a first-time BSI event associated with insomnia symptoms

|  |  |  |  |  |  |
| --- | --- | --- | --- | --- | --- |
|  | Person years | BSI events | Age adjusted* | Multivariable adjusted** |  |
|  |  |  | HR | HR | 95% CI |
| **Difficulty initiating sleep** |  |  |  |  |  |
| Never | 375,494 | 759 | 1.00 | 1.00 | Reference |
| Occasionally | 243,288 | 599 | 1.00 | 1.06 | 0.95-1.18 |
| Often/almost every night | 55,458 | 182 | 1.08 | 1.14 | 0.96-1.34 |
| Linear trend**** | 674,239 | 1,540 | 1.03 | 1.06 | 0.99-1.15 |
| **Difficulty maintaining sleep** |  |  |  |  |  |
| Never | 328,670 | 491 | 1.00 | 1.00 | Reference |
| Occasionally | 285,548 | 837 | 1.03 | 1.10 | 0.98-1.23 |
| Often/almost every night | 61,320 | 230 | 1.10 | 1.19 | 1.01-1.40 |
| Linear trend**** | 675,537 | 1,558 | 1.04 | 1.09 | 1.01-1.18 |
| **Feeling of nonrestorative sleep\*\*\*** |  |  |  |  |  |
| Never, few times a year | 408,694 | 534 | 1.00 | 1.00 | Reference |
| 1-2 Times per month | 98,093 | 166 | 1.10 | 1.15 | 0.96-1.37 |
| Once a week/more than once a week | 88,069 | 198 | 1.21 | 1.23 | 1.04-1.46 |
| Linear trend**** | 594,855 | 898 | 1.10 | 1.11 | 1.03-1.21 |
| **Cumulative insomnia symptoms\*\*\*** |  |  |  |  |  |
| 0 | 479,449 | 640 | 1.00 | 1.00 | Reference |
| 1 | 57,854 | 117 | 1.12 | 1.13 | 0.93-1.38 |
| 2 | 35,241 | 75 | 1.20 | 1.20 | 0.94-1.53 |
| 3 | 16,512 | 50 | 1.40 | 1.39 | 1.04-1.87 |
| Linear trend**** | 589,055 | 882 | 1.11 | 1.11 | 1.03-1.20 |
| *Age adjusted: adjusted for age (as a timescale).<br>**Multivariable adjusted: adjusted for age (as a timescale), sex (male, female), marital status (married/partner, separated/divorced/widower or never married), education (<10 years, 10–12 years, >12 years), BMI (< 18.5, 18.5–24.9, 25–29.9, 30.0–34.9, 35.0–39.9 and ≥40.0 kg/m<sup>2</sup>), smoking (never, former, current), alcohol consumption (abstainer, light drinker, moderate drinker, heavy drinker), physical activity (inactive, light activity, moderate activity, vigorous activity).<br>***Analysis restricted to age group 20 to 69 years (n=44,671).<br>**** Estimates when entering the insomnia categories (0–3) were entered as a continuous variable in the regression model.<br>Abbreviations: HR – hazard ratio, CI – confidence interval, BSI – bloodstream infection, BMI=body mass index (kg/m<sup>2</sup>). |  |  |  |  |  |

**Table 3.**
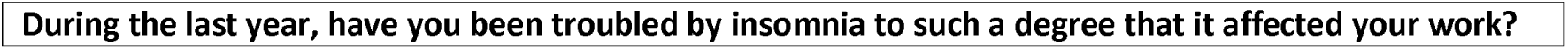
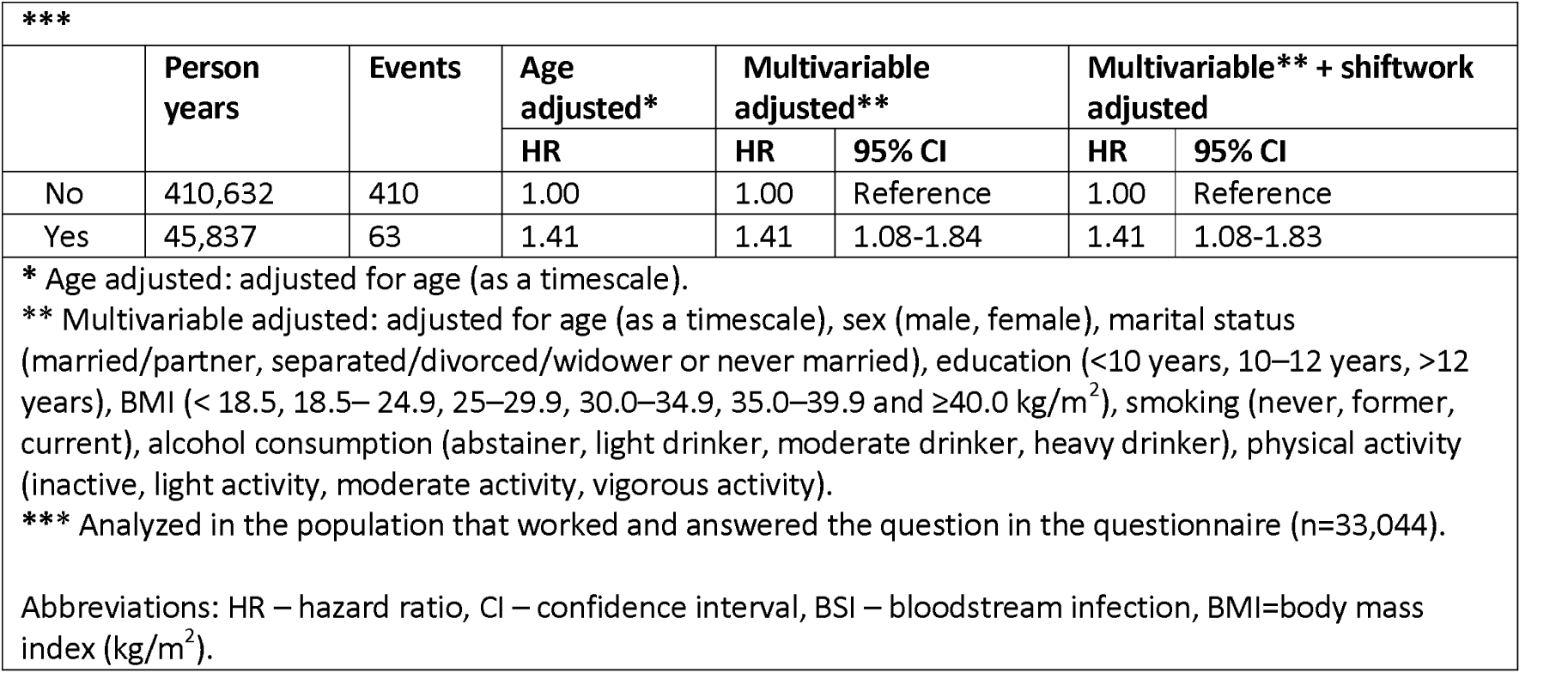
Risk of first-time BSI event related to insomnia affecting work performance

| *** |  |  |  |  |  |  |  |
| --- | --- | --- | --- | --- | --- | --- | --- |
|  | Person years | Events | Age adjusted* | Multivariable adjusted** |  | Multivariable** + shiftwork adjusted |  |
|  |  |  | HR | HR | 95% CI | HR | 95% CI |
| No | 410,632 | 410 | 1.00 | 1.00 | Reference | 1.00 | Reference |
| Yes | 45,837 | 63 | 1.41 | 1.41 | 1.08-1.84 | 1.41 | 1.08-1.83 |
| * Age adjusted: adjusted for age (as a timescale).<br>** Multivariable adjusted: adjusted for age (as a timescale), sex (male, female), marital status (married/partner, separated/divorced/widower or never married), education (<10 years, 10–12 years, >12 years), BMI (< 18.5, 18.5– 24.9, 25–29.9, 30.0–34.9, 35.0–39.9 and ≥40.0 kg/m<sup>2</sup>), smoking (never, former, current), alcohol consumption (abstainer, light drinker, moderate drinker, heavy drinker), physical activity (inactive, light activity, moderate activity, vigorous activity).<br>*** Analyzed in the population that worked and answered the question in the questionnaire (n=33,044).<br>Abbreviations: HR – hazard ratio, CI – confidence interval, BSI – bloodstream infection, BMI=body mass index (kg/m<sup>2</sup>). |  |  |  |  |  |  |  |

In sensitivity analyses, adjustment for shift work and doing our analysis in complete case had no effect on the results (data not shown). When adjusting for potentially mediating factors (comorbid conditions and HADS anxiety and depression scores) of the associations between insomnia symptoms and risk of BSI the HRs were somewhat attenuated (Supplementary Table 1).

Supplementary Table 2 shows the risk of BSI by specific bacteria. Regarding *E*.*coli* BSI, in analysis of linear trend, the HRs were 1.18 (95% CI 1.04–1.34) for “difficulties maintaining sleep”, and 1.12 (95% CI 1.00–1.27) for “difficulties initiating sleep”. This was not observed in analysis for linear trend for cumulative insomnia symptoms or for any insomnia symptoms related to *S. pneumonia or S. aureus*. For *S. pneumonia*, the HR pointed towards increased risk when having difficulties maintaining sleep and having a feeling of non-restorative sleep often or almost every night, but confidence intervals were also in line with no difference.

Of the total 1,579 participants experiencing BSI, 283 (18.0%) suffered BSI related mortality. Table 4 shows HRs of BSI related mortality according to categories of insomnia symptoms. The risk estimates for BSI related mortality associated with insomnia symptoms were similar to the estimates observed for first-time BSI. For example, compared to the reference category, the HRs for those having three cumulative insomnia symptoms was 1.45 (95% CI 0.72–2.93). However, due to relatively few BSI related deaths confidence intervals are wide and inconclusive.

**Table 4.** Risk of a BSI-related death associated with insomnia symptoms.

| **Table 4.** Risk of a BSI-related death associated with insomnia symptoms. |  |  |  |  |  |
| --- | --- | --- | --- | --- | --- |
|  | Person<br>years | Events | Age adjusted* | Multivariable<br>adjusted** |  |
|  |  |  | HR | HR | 95% CI |
| **Difficulty initiating**<br>**sleep** |  |  |  |  |  |
| Never | 375,494 | 138 | 1.00 | 1.00 | Reference |
| Occasionally | 243,288 | 109 | 0.96 | 1.04 | 0.80-1.34 |
| Often/almost every<br>night | 55,458 | 26 | 0.77 | 0.85 | 0.55-1.31 |
| Linear trend**** | 674,239 | 273 | 0.90 | 0.96 | 0.80-1.15 |
| **Difficulty maintaining**<br>**sleep** |  |  |  |  |  |
| Never | 328,670 | 82 | 1.00 | 1.00 | Reference |
| Occasionally | 285,548 | 152 | 0.94 | 1.06 | 0.81-1.40 |
| Often/almost every<br>night | 61,320 | 47 | 1.08 | 1.29 | 0.89-1.86 |
| Linear trend**** | 675,537 | 281 | 1.03 | 1.12 | 0.94-1.35 |
| **Feeling of**<br>**nonrestorative**<br>**sleep\*\*\*** |  |  |  |  |  |
| Never, few times a<br>year | 408,694 | 75 | 1.00 | 1.00 | Reference |
| 1-2 Times per month | 98,093 | 21 | 0.95 | 0.99 | 0.61-1.62 |
| Once a week/more<br>than once a week | 88,069 | 35 | 1.37 | 1.40 | 0.92-2.12 |
| Linear trend**** | 594,855 | 131 | 1.15 | 1.17 | 0.95-1.44 |
| **Cumulative insomnia**<br>**symptoms\*\*\*** |  |  |  |  |  |
| 0 | 479,449 | 91 | 1.00 | 1.00 | Reference |
| 1 | 57,854 | 21 | 1.28 | 1.30 | 0.80-2.10 |
| 2 | 35,241 | 9 | 0.94 | 0.93 | 0.47-1.86 |
| 3 | 16,512 | 9 | 1.55 | 1.45 | 0.72-2.93 |
| Linear trend**** | 589,055 | 130 | 1.11 | 1.09 | 0.90-1.33 |
| * Age adjusted: adjusted for age (as a timescale).<br>** Multivariable adjusted: adjusted for age (as a timescale), sex (male, female), marital status (married/partner, separated/divorced/widower or never married), education (<10 years, 10–12 years, >12 years), BMI (< 18.5, 18.5– 24.9, 25–29.9, 30.0–34.9, 35.0–39.9 and ≥40.0 kg/m<sup>2</sup>), smoking (never, former, current), alcohol consumption (abstainer, light drinker, moderate drinker, heavy drinker), physical activity (inactive, light activity, moderate activity, vigorous activity).<br>*** Analysis performed in the age group 20 to 69 (n=44,671)<br>**** Estimates when entering the insomnia categories (0–3) were entered as a continuous variable in the regression model.<br>Abbreviations: HR – hazard ratio, CI – confidence interval, BSI – bloodstream infection, BMI=body mass index (kg/m<sup>2</sup>). |  |  |  |  |  |

In the sensitivity analysis examining the effect of light exposure at the time of reported insomnia symptoms on BSI outcomes, we observed no appreciable changes in our results (data not shown). When we analyzed risk of BSI in the population not using sedatives on a weekly basis, the risk was higher for participants with three cumulative insomnia symptoms than for the total population, with a 56% (95% CI 6%–131%) increased risk (Supplementary Table 3). We observed no notable differences in risk estimates when investigating the influence of reverse causation (Supplementary Table 4).

When assessing effect modification by age (±60 years) and sex, we found no evidence of interaction in any of the analysis (all p>0.2).

## Discussion

In this study prospective population-based study of 53,536 individuals we observed weak positive associations between all measures of insomnia symptoms and risk of BSI. People who reported three cumulative insomnia symptoms had 40% increased risk of future BSI compared to participants having no insomnia symptoms. Among the working population, those who reported that their insomnia affected their ability to work were also at 40% increased risk of BSI. The results for BSI related mortality were inconclusive. Similar to our study, a previous retrospective study following 24,173 Taiwanese participants for 10 years, found 143% increased risk of pneumonia among participants with ICD-9 diagnosis of insomnia, compared with participants without this diagnosis (Lin et al., 2018). As Lin et al. investigated pneumonia and we investigated bloodstream infection, the higher effect estimates could be due to different outcomes and adjustments for less confounders than in our study. However, as we found evidence of a dose-response relationship between severity of insomnia symptoms and BSI risk we speculate that the higher estimates in Lin et al. reflect that an insomnia diagnosis is a higher indicator of the exposure than self-report.

Results from the bacteria-specific analyses point in the direction of insomnia symptoms being an increased risk for future BSI of E. coli. These results regarding E. coli coincide with previous studies having found sleep deprived rabbits to be more likely to get a positive blood culture, have higher morbidity and mortality when exposed to E. coli, than rabbits with enhanced sleep (Toth, Tolley, & Krueger, 1993). However, as Tooth et al (1993) found the risk for BSI in rabbits also related to *S. aureus*, it seems most plausible that our negative results for other bacteria are due to few events and thus lower precision.

The risk estimates for BSI related mortality with the insomnia items pointed towards a 40% increased risk but with imprecise estimates due to few events. However, a meta-analysis that found short sleep duration resulted in a higher all-cause mortality can give support to the plausibility of our effect estimates (Gallicchio & Kalesan, 2009), especially as short sleep duration might be similar to our items regarding difficulty initiating sleep or difficulty maintain sleep.

Insomnia posed similar risk of BSI with seasonal variations in light exposure in our study. This could be explained by insomnia being a stable trait, not being affected by seasonal variation neither at 63°–65° degrees north where the HUNT study is performed nor further north near the polar circle (Shochat et al., 2019; Sivertsen, Friborg, Pallesen, Vedaa, & Hopstock, 2020). When analyzing risk of BSI in the population not using sedatives, risk was 50% among participants having all three cumulative insomnia symptoms, which is a higher than the 40% risk with three cumulative insomnia symptoms in the total population. Interpreting these results is difficult as we do not know whether the participants answered how their sleep is with or without sedatives. Our study should not be taken into account of a reduced risk of BSI with sleeping medication without further investigations, as previous studies have shown that some sleep medications show increased risk of infection with insomnia (Huang et al., 2014; Kripke, 2016).

Chronic diseases may be co-present with sleeping disturbances (Besedovsky, Lange, & Haack, 2019b) and could therefore also be a confounding factor associated with both insomnia and BSI or other infections. In the studies most comparable to ours, they either lacked information regarding comorbid conditions (Cohen et al., 2009; Prather et al., 2015), was limited to a female population (Patel et al., 2012) or they were not able to adjust for other potential important confounders (Lin et al., 2018; Prather & Leung, 2016). In contrast our study is not limited by sex and includes both presumably healthy participants and participants with comorbid conditions, and we are also able to adjust for important lifestyle factors. We tested for reverse causality by excluding the first five years of follow-up and found no notable changes in risk.

### Strength and limitations

Major strengths of our study include the large population with a prospective design and minimal loss to follow-up. As most large population-based studies looking at the association between insomnia and future risk of infection have used self-reported subjective symptoms of the outcome (Patel et al., 2012; Prather & Leung, 2016), a strength of our study is the use of a confirmed severe invasive infection as our outcome. By using a positive blood culture, we have a more objective measure of serious infection than most other large studies. As an example, in pneumonia, the phenotype of the infectious agent is often unknown, and the heterogeneity of the disease is known to be large (Mackenzie, 2016). This could make comparing between studies difficult. Due to the rich baseline information in HUNT2, we were able to adjust analyses for important lifestyle factors (smoking, alcohol consumption, BMI and activity level) that were not available in previous studies (Lin et al., 2018). As with any observational study, however, limitations include a risk of residual confounding (Rothman, Greenland, & Lash, 2008) Further, this study cannot establish the causal pathways between insomnia and BSI. Based on our apriori subject matter knowledge, our most reliable confounders are sex and age, while lifestyle factors, comorbidities, anxiety and depression symptoms have more unclear pathways. Insomnia could both be a result of or be worsened by poor somatic health, and it could worsen both mental and physical health (Besedovsky et al., 2019b; Grandner, 2020). Lifestyle factors are likely to affect sleeping patterns, and symptoms of insomnia would also affect the ability to perform normal daily activities. As our analysis were directed by our hypothesis, we therefore assume model 2 to reflect the best interpretation of our analyses as adjustments for age, sex and important lifestyle factors are included.

Limitations include the lack of DSM-IV or -V specific diagnostic criteria for insomnia, as the questionnaire ask of symptoms the last month and not for longer durations. However, it is known that prevalence estimates of insomnia are lower in studies using DSM-IV compared to studies not assessing the severity and frequency of sleeping problems as strictly (Ohayon & Partinen, 2002). Due to this, a significant amount of people with sleep difficulties may not fit into the diagnostic criteria (Ohayon & Reynolds, 2009). As Edinger et al. point out, the use of a strict criteria for prevalence may cause an underestimation of the amount of participants with clinically important sleep problems (Edinger & Means, 2005). We also do not have information on whether the participants’ sleeping pattern is a temporary situation or their regular habits, as insomnia was only evaluated at the start of the study. And last, no evaluation of sleep apnea syndrome was available.

## Conclusion

This is the first human-based study to date regarding insomnia as a risk factor for future BSI and BSI related mortality. We found evidence of a weak positive increased risk associated of first-time BSI with all insomnia symptoms as well as moderately increased risk with cumulative insomnia symptoms and insomnia to such a degree that it affected work performance. More research is needed to understand the underlying mechanisms causing increased risk of BSI with increasing symptoms of insomnia. As BSI is a global concern, addressing modifiable risk factors such as insomnia is of utmost importance to reduce the burden of BSI. By understanding this causal relationship, we could possibly prevent BSI in the future by addressing insomnia as a preventable risk factor.

## Data Availability

All data produced in the present study are available upon reasonable request to the authors.

## Acknowledgments

We would like to thank the Trøndelag Health Study (HUNT), a collaboration of the HUNT Research Center (Faculty of Medicine and Health Sciences, NTNU, Norwegian University of Science and Technology), Trøndelag County Council, Central Norway Regional Health Authority and the Norwegian Institute of Public Health. In addition, we also thank the microbiology departments at the hospitals for providing microbial data and the Department for Research at Nord-Trøndelag Hospital Trust for assistance with data linkage.

---

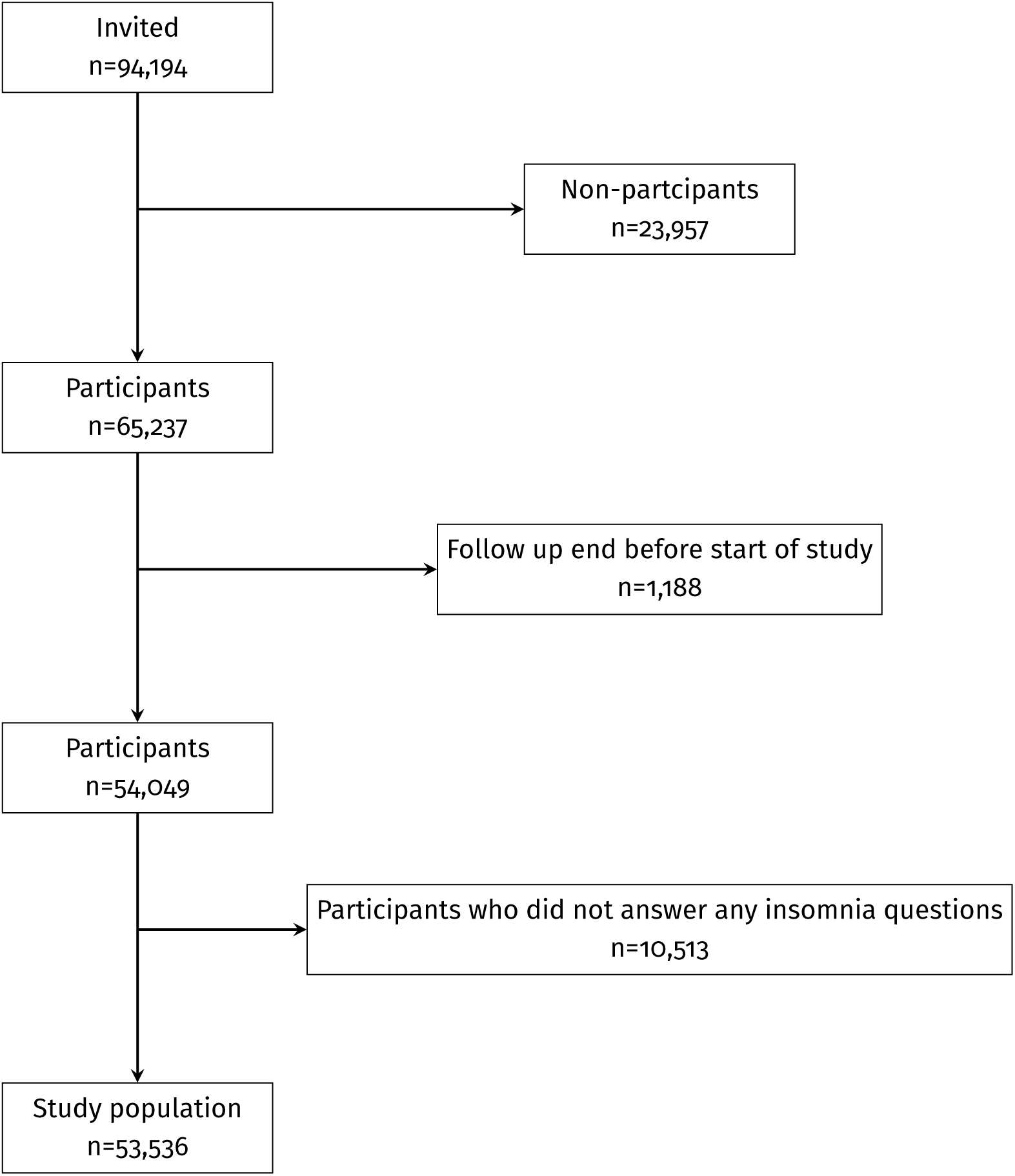

