## Supplemental Tables for "Insomnia symptoms and risk of bloodstream infections: prospective data from the prospective population-based HUNT Study, Norway"

**Supplementary Table 1.** Risk of first-time BSI event, adjusted for comorbidities and HADS depression and anxiety score, associated with insomnia symptoms.

| | Person-years | Events | Age adjusted* | | Multivariable adjusted** | | Multivariable adjusted + comorbidities*** | | Multivariable adjusted + comorbidities and HADS score**** | |
|---|---|---|---|---|---|---|---|---|---|---|
| | | | HR | 95% CI | HR | 95% CI | HR | 95% CI | HR | 95% CI |
| **Difficulty initiating sleep** | | | | | | | | | | |
| Never | 375,494 | 759 | 1.00 | Reference | 1.00 | Reference | 1.00 | Reference | 1.00 | Reference |
| Occasionally | 243,288 | 599 | 1.00 | 0.90-1.11 | 1.06 | 0.95-1.18 | 1.03 | 0.93-1.15 | 1.02 | 0.91-1.14 |
| Often/almost every night | 55,458 | 182 | 1.08 | 0.92-1.27 | 1.14 | 0.96-1.34 | 1.07 | 0.90-1.26 | 1.04 | 0.87-1.24 |
| Linear trend****** | 674,239 | 1,540 | 1.03 | 0.95-1.11 | 1.06 | 0.99-1.15 | 1.03 | 0.96-1.11 | 1.02 | 0.94-1.11 |
| **Difficulty maintaining sleep** | | | | | | | | | | |
| Never | 328,670 | 491 | 1.00 | Reference | 1.00 | Reference | 1.00 | Reference | 1.00 | Reference |
| Occasionally | 285,548 | 837 | 1.03 | 0.92-1.15 | 1.10 | 0.98-1.23 | 1.08 | 0.96-1.21 | 1.07 | 0.95-1.21 |
| Often/almost every night | 61,320 | 230 | 1.10 | 0.94-1.29 | 1.19 | 1.01-1.40 | 1.12 | 0.95-1.32 | 1.10 | 0.93-1.31 |
| Linear trend****** | 675,537 | 1,558 | 1.04 | 0.97 | 1.09 | 1.01-1.18 | 1.06 | 0.98-1.15 | 1.05 | 0.97-1.14 |
| **Feeling of nonrestorative sleep***** | | | | | | | | | | |
| Never, few times a year | 408,694 | 534 | 1.00 | Reference | 1.00 | Reference | 1.00 | Reference | 1.00 | Reference |
| 1-2 Times per month | 98,093 | 166 | 1.10 | 0.93-1.31 | 1.15 | 0.96-1.37 | 1.13 | 0.95-1.34 | 1.11 | 0.93-1.33 |
| Once a week/ more than once a week | 88,069 | 198 | 1.21 | 1.02-1.42 | 1.23 | 1.04-1.46 | 1.17 | 0.99-1.39 | 1.15 | 0.96-1.37 |
| Linear trend****** | 594,855 | 898 | 1.10 | 1.01-1.19 | 1.11 | 1.03-1.21 | 1.09 | 1.00-1.18 | 1.07 | 0.98-1.17 |
| **Cumulative insomnia symptoms***** | | | | | | | | | | |
| 0 | 479,449 | 640 | 1.00 | Reference | 1.00 | Reference | 1.00 | Reference | 1.00 | Reference |
| 1 | 57,854 | 117 | 1.12 | 0.92-1.36 | 1.13 | 0.93-1.38 | 1.10 | 0.90-1.34 | 1.08 | 0.88-1.32 |
| 2 | 35,241 | 75 | 1.20 | 0.95-1.53 | 1.20 | 0.94-1.53 | 1.15 | 0.90-1.46 | 1.11 | 0.86-1.42 |

| | | | | | | | | | | |
|---|---|---|---|---|---|---|---|---|---|---|
| 3 | 16,512 | 50 | 1.40 | 1.05-1.87 | 1.39 | 1.04-1.87 | 1.30 | 0.97-1.74 | 1.23 | 0.91-1.68 |
| Linear trend[******] | 589,055 | 882 | 1.11 | 1.03-1.20 | 1.11 | 1.03-1.20 | 1.08 | 1.00-1.17 | 1.07 | 0.98-1.16 |

* Age adjusted: Adjusted for age (as a timescale).

** Multivariable adjusted:  (as a timescale) and sex (male, female), marital status (married/partner, separated/divorced/widower or never married), education (<10 years, 10–12 years, >12 years), BMI (< 18.5, 18.5– 24.9, 25–29.9, 30.0–34.9, 35.0–39.9 and ≥40.0 kg/m2), smoking (never, former, current), alcohol consumption (abstainer, light drinker, moderate drinker, heavy drinker), physical activity (inactive, light activity, moderate activity, vigorous activity).

*** Multivariable + comorbidities adjusted: Multivariable adjusted and adjusted for comorbid conditions (chronic vascular disease, chronic kidney disease, lung disease, cancer, diabetes and rheumatic disease).

**** Multivariable + comorbidities and HADS score adjusted: Multivariable and comorbidities adjusted and adjusted for HADS anxiety and depression score.

***** Analysis performed in the age group 20 to 69 (n=44,671).

****** Estimates when entering the insomnia categories (0–3) were entered as a continuous variable in the regression model

Abbreviations: HR – hazard ratio, CI – confidence interval, BSI – bloodstream infection, BMI=body mass index (kg/m2)

**Supplementary Table 2**. Risk of first-time event BSI event specified by bacteria.

| | | E. Coli | | | S. Pneumonia | | | S. Aureus | | |
|---|---|---|---|---|---|---|---|---|---|---|
| | Person-years | Events | Multivariable adjusted* | | Events | Multivariable adjusted* | | Events | Multivariably adjusted* | |
| | | | HR | 95% CI | | HR | 95% CI | | HR | 95% CI |
| **Difficulty initiating sleep** | | | | | | | | | | |
| Never | 375,494 | 257 | 1.00 | Reference | 105 | 1.00 | Reference | 97 | 1.00 | Reference |
| Occasionally | 243,288 | 242 | 1.18 | 0.99-1.41 | 74 | 0.95 | 0.71-1.29 | 63 | 0.91 | 0.66-1.25 |
| Often/almost every night | 55,458 | 74 | 1.22 | 0.93-1.59 | 20 | 0.93 | 0.57-1.51 | 20 | 1.01 | 0.62-1.66 |
| Linear trend*** | 674,239 | 573 | 1.12 | 1.00-1.27 | 199 | 0.96 | 0.77-1.19 | 180 | 0.97 | 0.78-1.21 |
| **Difficulty maintaining sleep** | | | | | | | | | | |
| Never | 328,670 | 150 | 1.00 | Reference | 69 | 1.00 | Reference | 62 | 1.00 | Reference |
| Occasionally | 285,548 | 345 | 1.36 | 1.01-1.65 | 107 | 1.13 | 0.82-1.55 | 96 | 1.02 | 0.73-1.41 |
| Often/almost every night | 61,320 | 90 | 1.33 | 1.02-1.74 | 26 | 1.12 | 0.70-1.78 | 22 | 0.92 | 0.56-1.52 |
| Linear trend*** | 675,537 | 585 | 1.18 | 1.04-1.34 | 202 | 1.08 | 0.86-1.34 | 180 | 0.97 | 0.77-1.23 |
| **Feeling of nonrestorative sleep**** | | | | | | | | | | |
| Never, few times a year | 408,694 | 179 | 1.00 | Reference | 77 | 1.00 | Reference | 65 | 1.00 | Reference |
| 1-2 Times per month | 98,093 | 61 | 1.16 | 0.86-1.55 | 25 | 1.23 | 0.78-1.94 | 13 | 0.78 | 0.43-1.43 |
| Once a week/ more than once a week | 88,069 | 76 | 1.21 | 0.92-1.60 | 28 | 1.29 | 0.83-2.02 | 22 | 1.12 | 0.68-1.84 |
| Linear trend*** | 594,855 | 316 | 1.11 | 0.97-1.27 | 130 | 1.15 | 0.92-1.42 | 100 | 1.03 | 0.80-1.32 |
| **Cumulative insomnia symptoms**** | | | | | | | | | | |
| 0 | 479,449 | 214 | 1.00 | Reference | 97 | 1.00 | Reference | 75 | 1.00 | Reference |
| 1 | 57,854 | 49 | 1.31 | 0.95-1.79 | 18 | 1.22 | 0.73-2.02 | 10 | 0.83 | 0.43-1.61 |
| 2 | 35,241 | 28 | 1.20 | 0.80-1.79 | 10 | 1.07 | 0.55-2.06 | 6 | 0.80 | 0.35-1.86 |
| 3 | 16,512 | 19 | 1.35 | 0.84-2.18 | 5 | 0.97 | 0.39-2.42 | 7 | 1.57 | 0.71-3.49 |

| | | | | | | | | | |
|---|---|---|---|---|---|---|---|---|---|
| Linear trend*** | 589,055 | 310 | 1.12 | 0.99-1.27 | 130 | 1.03 | 0.83-1.28 | 98 | 1.05 | 0.82-1.33 |

* Multivariable adjusted: adjusted for age (as a timescale), sex (male, female), marital status (married/partner, separated/divorced/widower or never married), education (<10 years, 10–12 years, >12 years), BMI (< 18.5, 18.5– 24.9, 25–29.9, 30.0–34.9, 35.0–39.9 and ≥40.0 kg/m2), smoking (never, former, current), alcohol consumption (abstainer, light drinker, moderate drinker, heavy drinker), physical activity (inactive, light activity, moderate activity, vigorous activity).

** Analysis restricted to age group 20 to 69 years (n=44,671).

*** Estimates when entering the insomnia categories (0–3) were entered as a continuous variable in the regression model.

Abbreviations: HR – hazard ratio, CI – confidence interval, BSI – bloodstream infection, BMI=body mass index (kg/m2).

**Supplementary Table 3.** Risk of first-time BSI event in a population reportion to not use sedatives on a regular basis.

| | Person-yeras | Events | Age adjusted* | | Multivariable adjusted** | |
|---|---|---|---|---|---|---|
| | | | HR | 95% CI | HR | 95% CI |
| **Difficulty initiating sleep** | | | | | | |
| Never | 330,409 | 655 | 1.00 | Reference | 1.00 | Reference |
| Occasionally | 203,945 | 459 | 0.97 | 0.86-1.10 | 1.04 | 0.92-1.17 |
| Often/almost every night | 31,950 | 81 | 1.18 | 0.93-1.48 | 1.24 | 0.98-1.56 |
| Linear trend**** | 566,304 | 1,195 | 1.03 | 0.94-1.13 | 1.08 | 0.98-1.18 |
| **Difficulty maintaining sleep** | | | | | | |
| Never | 289,799 | 428 | 1.00 | Reference | 1.00 | Reference |
| Occasionally | 235,679 | 638 | 0.99 | 0.87-1.12 | 1.07 | 0.94-1.21 |
| Often/almost every night | 41,249 | 142 | 1.14 | 0.94-1.39 | 1.24 | 1.02-1.51 |
| Linear trend**** | 566,727 | 1,208 | 1.05 | 0.95-1.15 | 1.10 | 1.01-1.21 |
| **Feeling of nonrestorative sleep***** | | | | | | |
| Never, few times a year | 361,521 | 467 | 1.00 | Reference | 1.00 | Reference |
| 1-2 Times per month | 84,691 | 136 | 1.07 | 0.89-1.30 | 1.16 | 0.96-1.41 |
| Once a week/ more than once a week | 62,987 | 123 | 1.14 | 0.93-1.39 | 1.23 | 1.02-1.48 |
| Linear trend**** | 509,199 | 726 | 1.07 | 0.97-1.17 | 1.10 | 1.00-1.21 |
| **Cumulative insomnia symptoms***** | | | | | | |
| 0 | 423,884 | 558 | 1.00 | Reference | 1.00 | Reference |
| 1 | 47,4598 | 88 | 1.06 | 0.85-1.33 | 1.09 | 0.87-1.37 |
| 2 | 24,856 | 44 | 1.10 | 0.81-1.49 | 1.12 | 0.82-1.53 |
| 3 | 9,215 | 27 | 1.53 | 1.04-2.25 | 1.56 | 1.06-2.31 |
| Linear trend**** | 505,554 | 717 | 1.10 | 1.00-1.21 | 1.11 | 1.01-1.23 |

* Age adjusted: Adjusted for age (as a timescale).

** Multivariable adjusted: Age adjusted (as a timescale) and sex (male, female), marital status (married/partner, separated/divorced/widower or never married), education (<10 years, 10–12 years, >12 years), BMI (< 18.5, 18.5– 24.9, 25–29.9, 30.0–34.9, 35.0–39.9 and ≥40.0 kg/m$^2$), smoking (never, former, current), alcohol consumption (abstainer, light drinker, moderate drinker, heavy drinker), physical activity (inactive, light activity, moderate activity, vigorous activity).

*** Analysis performed in the age group 20 to 69 (n=43.198).

**** Estimates when entering the insomnia categories (0–3) were entered as a continuous variable in the regression model

Abbreviations: HR – hazard ratio, CI – confidence interval, BSI – bloodstream infection, BMI=body mass index (kg/m$^2$).

**Supplementary Table 4** – Risk of first-time BSI event adjusted for comorbidities excluding the first five years of follow up.

| | Person-years | Events | Age adjusted* | | Multivariable adjusted** | |
|---|---|---|---|---|---|---|
| | | | HR | 95% CI | HR | 95% CI |
| **Difficulty initiating sleep** | | | | | | |
| Never | 258,912 | 595 | 1.00 | Reference | 1.00 | Reference |
| Occasionally | 166,953 | 475 | 1.00 | 0.89-1.13 | 1.07 | 0.95-1.21 |
| Often/almost every night | 37,364 | 131 | 1.02 | 0.84-1.23 | 1.08 | 0.89-1.31 |
| Linear trend**** | 463,229 | 1,201 | 1.01 | 0.92-1.09 | 1.05 | 0.96-1.14 |
| **Difficulty maintaining sleep** | | | | | | |
| Never | 227,416 | 390 | 1.00 | Reference | 1.00 | Reference |
| Occasionally | 195,216 | 650 | 1.01 | 0.89-1.15 | 1.07 | 0.94-1.22 |
| Often/almost every night | 41,396 | 176 | 1.08 | 0.90-1.29 | 1.16 | 0.97-1.39 |
| Linear trend**** | 464,027 | 1,216 | 1.03 | 0.94-1.13 | 1.08 | 0.98-1.18 |
| **Feeling of nonrestorative sleep*** ** | | | | | | |
| Never, few times a year | 286,385 | 440 | 1.00 | Reference | 1.00 | Reference |
| 1-2 Times per month | 68,378 | 138 | 1.11 | 0.91-1.34 | 1.16 | 0.96-1.41 |
| Once a week/ more than once a week | 61,435 | 162 | 1.19 | 0.99-1.43 | 1.23 | 1.02-1.48 |
| Linear trend**** | 416,435 | 740 | 1.09 | 1.00-1.20 | 1.11 | 1.02-1.22 |
| **Cumulative insomnia symptoms*** ** | | | | | | |
| 0 | 335,735 | 528 | 1.00 | Reference | 1.00 | Reference |
| 1 | 40,458 | 98 | 1.13 | 0.91-1.40 | 1.15 | 0.92-1.43 |
| 2 | 24,567 | 57 | 1.10 | 0.84-1.45 | 1.11 | 0.85-1.47 |
| 3 | 11,498 | 42 | 1.43 | 1.04-1.96 | 1.42 | 1.03-1.96 |
| Linear trend**** | 412,197 | 725 | 1.10 | 1.01-1.20 | 1.10 | 1.01-1.20 |

* Age adjusted: Adjusted for age (as a timescale).

**Multivariable adjusted: Adjusted for age (as a timescale), sex (male, female), marital status (married/partner, separated/divorced/widower or never married), education (<10 years, 10-12 years, >12 years), BMI (< 18.5, 18.5– 24.9, 25–29.9, 30.0–34.9, 35.0–39.9 and ≥40.0 kg/m$^2$), smoking (never, former, current), alcohol consumption (abstainer, light drinker, moderate drinker, heavy drinker), physical activity (inactive, light activity, moderate activity, vigorous activity).

***Analysis performed in the age group 20 to 69 (n=44,671).

****Estimates when entering the insomnia categories as a continues variable in the regression model.

Abbreviations: HR – Hazard ratio, CI – confidence interval, BSI – Bloodstream infection, BMI=Body mass index (kg/m$^2$).

**Supplementary table 5** – Risk of first-time BSI event  specified by sex.

| | Female | | | | Male | | | |
|---|---|---|---|---|---|---|---|---|
| | Person-years | Events | Multivariable adjusted* | | Person-years | Events | Multivariable adjusted* | |
| | | | HR | 95% CI | | | HR | 95% CI |
| **Difficulty initiating sleep** | | | | | | | | |
| Never | 192.328 | 295 | 1.00 | Reference | 183.165 | 464 | 1.00 | Reference |
| Occasionally | 141.929 | 328 | 1.14 | 0.97-1.33 | 101.359 | 271 | 0.99 | 0.85-1.15 |
| Often/almost every night | 37.560 | 113 | 1.09 | 0.87-1.35 | 17.858 | 69 | 1.24 | 0.96-1.61 |
| Linear trend*** | 371.857 | 736 | 1.06 | 0.96-1.18 | 302.382 | 804 | 1.06 | 0.95-1.18 |
| **Difficulty maintaining sleep** | | | | | | | | |
| Never | 171.007 | 193 | 1.00 | Reference | 157.662 | 298 | 1.00 | Reference |
| Occasionally | 162.616 | 414 | 1.16 | 0.97-1.38 | 122.931 | 423 | 1.06 | 0.91-1.24 |
| Often/almost every night | 39.386 | 141 | 1.30 | 1.04-1.64 | 21.934 | 89 | 1.08 | 0.85-1.37 |
| Linear trend*** | 373.010 | 748 | 1.14 | 1.02-1.28 | 302.528 | 810 | 1.05 | 0.94-1.17 |
| **Feeling of nonrestorative sleep**** | | | | | | | | |
| Never, few times a year | 210.957 | 222 | 1.00 | Reference | 197.736 | 312 | 1.00 | Reference |
| 1-2 Times per month | 55.578 | 90 | 1.22 | 0.95-1.56 | 42.515 | 76 | 1.08 | 0.84-1.39 |
| Once a week/ more than once a week | 58.220 | 127 | 1.25 | 1.00-1.56 | 29.849 | 71 | 1.20 | 0.93-1.56 |
| Linear trend** | 324.755 | 439 | 1.12 | 1.01-1.25 | 270.100 | 459 | 1.09 | 0.97-1.24 |
| **Cumulative insomnia symptoms**** | | | | | | | | |
| 0 | 251.477 | 277 | 1.00 | Reference | 227.971 | 363 | 1.00 | Reference |
| 1 | 34.520 | 70 | 1.29 | 0.99-1.68 | 23.334 | 47 | 0.96 | 0.71-1.30 |
| 2 | 22.908 | 46 | 1.20 | 0.88-1.65 | 12.333 | 29 | 1.20 | 0.82-1.76 |
| 3 | 11.537 | 34 | 1.41 | 0.98-2.03 | 4.975 | 16 | 1.39 | 0.84-2.29 |
| Linear trend*** | 320.442 | 427 | 1.12 | 1.02-1.24 | 268.614 | 455 | 1.09 | 0.96-1.23 |

* Multivariable adjusted: Adjusted for age (as a timescale), sex (male, female), marital status (married/partner, separated/divorced/widower or never married), education (<10 years, 10-12 years, >12 years), BMI (< 18.5, 18.5– 24.9, 25–29.9, 30.0–34.9, 35.0–39.9 and ≥40.0 kg/m$^2$), smoking (never, former, current), alcohol consumption (abstainer, light drinker, moderate drinker, heavy drinker), physical activity (inactive, light activity, moderate activity, vigorous activity).

** Analysis performed in the age group 20 to 69 (n=44,671).

*** Estimates when entering the insomnia categories as a continues variable in the regression model.

Abbreviations: HR – Hazard ratio, CI – confidence interval, BSI – Bloodstream infection, BMI=Body mass index (kg/m$^2$).
